# Modelling the impact of larviciding as a supplementary malaria vector control intervention in rural south-eastern Tanzania: A district-level simulation study

**DOI:** 10.1101/2025.11.13.25340176

**Authors:** GloriaSalome Shirima, Emma L Fairbanks, Gavana Tegemeo, Gerald Kiwelu, Ismail Nambunga, Yeromin P Mlacha, Silas Mirau, Prosper Chaki, Nakul Chitnis, Samson Kiware

**Affiliations:** Environmental Health and Ecological Science Department, Ifakara Health Institute, P.O. Box 78373, Ifakara, Tanzania; Computational and Communication Science and Engineering, The Nelson Mandela African Institution of Science and Technology, (NM-AIST), P.O. Box 447, Tengeru, Arusha, Tanzania; Disease Modelling, Swiss Tropical and Public Health Institute, Kreuzstrasse 2, 4123, Allschwil, Switzerland; Department of Mathematics, University of Manchester, Manchester, UK; University of Basel, Petersplatz 1, 4001, Basel, Switzerland

**Keywords:** Larviciding, ITNs, VCOM, malaria transmission, district-level modeling

## Abstract

Combining larviciding with insecticide treated nets (ITNs) can reduce malaria transmission, but most modelling analyses use generalized scenarios rather than local contexts. In Tanzania and other countries, larviciding is increasingly being prioritized in national strategies, with growing advocacy for its broader implementation, to achieving sustained malaria reduction. District-specific modelling is therefore essential to capture variation in transmission ecology, seasonality, and varying coverage levels, providing evidence that is both rigorous and actionable for malaria control programs. The Vector Control Optimization Model (VCOM) was adapted and extended to incorporate local seasonality, simulating the impact of larviciding across a range of coverage levels combined with ITNs. The model was parameterized using district-level field-data on mosquito mortality collected before (2016-2017) and after (2019-2021) larviciding implementation. Mosquito mortality rates were estimated using Bayesian inference. Outcomes were evaluated specifically for *Anopheles gambiae* s.l. including annual entomological inoculation rates (EIR) and mosquito density. Sensitivity analysis explored the influence of key parameters driving transmission in this scenario study. The immature mosquito mortality rate due to larviciding is estimated to be 61% based on field data. VCOM simulation showed that, at 80%, ITNs coverage, larviciding substantially reduced mosquito densities and EIR. Specifically, combining ITNs at 80% and larviciding coverage ≥ 60% lowered EIR below 1, the threshold required to interrupt malaria transmission. Sensitivity analyses highlighted the high impact of targeting immature mosquitoes, suggesting larviciding can effectively complement ITNs to control vectors, including invasive species like *An. stephensi,* regardless of feeding preference, resting, and biting behaviors, which hinder the effectiveness of most vector control tools. This study provides local evidence that larviciding is an effective complement to ITNs for interrupting malaria transmission. Implementation should leverage innovative approaches, such as drones for precise mapping and targeted application of biological larvicides, to maximize coverage, and scalability for district-level malaria control and elimination.

## Background

Malaria, caused by the *Plasmodium* protozoan parasite and transmitted by a female anopheles’ mosquito, remains a significant global health threat (1). In 2023, there were an estimated 263 million cases globally, with the African region accounting for 89.7% of this burden (2). In Tanzania, malaria ranks among the top causes of mortality and morbidity; malaria case incidence increased by 5.6% from 121 in 2022 to 128 per 1000 of the population at risk (2–4). The Rufiji district, found in south-eastern Tanzania, is a recognized malaria hotspot within the country (5). The World Health Organization (WHO) recommends chemoprevention, case management, and vector control as core malaria interventions. However, the primary vector control tools, notably insecticide-treated nets (ITNs) and indoor residual spraying (IRS), are facing challenges. This is due to several factors, including insecticide resistance and changes in mosquito behavior, such as increased outdoor and early-evening biting, which reduce the effectiveness of these indoor-targeted tools (6). These challenges have contributed to a shortfall in achieving national and global targets such as the WHO’s 2030 goal to reduce malaria incidence and mortality by at least 90% from 2015 levels. As of 2023, globally, there were 60 cases per 1,000 people at risk and 13.7 deaths per 100,000 people at risk, compared to the targeted figures of 21.3 cases per 1,000 and 5.5 deaths per 100,000 people at risk (2, 7).

To address these limitations, the WHO promotes the Integrated Vector Management (IVM) strategy for sustainable malaria control and elimination (8). A combination of various interventions aims to target mosquitoes at multiple points in their lifecycle and in diverse settings, both indoors and outdoors. Key elements of IVM include evidence-based decision-making, the integration of non-chemical and chemical vector control methods, advocacy and social mobilization, and inter-sectoral collaboration and action. The key elements have recently been re-emphasized and endorsed by the World Health Assembly (WHA) in the form of pillars of action in the Global Vector Control Response framework for 2017–2030 (8). Innovative tools are needed to supplement existing ones rather than replace them. These include application of larviciding to affect the growth of immature mosquitoes (9), use of attractive toxic sugar baits (ATSBs) to lure and kill adult mosquitoes seeking sugar sources (10), treatment of cattle with endectocides to target mosquitoes that feed on livestock and reduce vector survival (11), house improvement and modification such as screening, sealing eaves, and using improved housing materials to minimize mosquito entry (12), and deployment of spatial repellents that create a protective barrier and reduce human–mosquito contact during outdoor activities (13). Further research is needed to understand the combined impact of these interventions in targeted areas. For instance, IVM programs incorporating larviciding with *Bacillus thuringiensis israelensis* (Bti) to the core vector interventions have been adopted in several African countries such as Kenya and Ethiopia contributing to significant malaria reductions (14, 15).

An alternative strategy to overcome the ITNs and IRS constraints is to focus on the larval phases of mosquitoes in their breeding areas, where they are densely populated, confined, and readily reachable (16). Larviciding is an intervention that aims at reducing the larval abundance, subsequently reducing the adult population by increasing the larval mortality. Larviciding process can include the use of chemicals, oils, or bio-larvicides (17). Bio-larvicides are currently advised for use, as they are environmentally friendly (18). Larviciding has shown a remarkable impact to complement ITNs and IRS targeting mosquitoes in the larval stage to interrupt malaria transmission (19), with different studies done around Sub-Saharan Africa showing that community-based involvement in the application of larviciding has brought a positive impact on the reduction of mosquito density (20–23). In Tanzania, larviciding has been added in the 2021-205 Malaria Strategic Plan as a supplementary vector control intervention alongside ITNs and IRS (5). There is growing advocacy for its broader implementation and its prioritization as one of the complementary interventions against vector control. Dar es Salaam, where larviciding was first implemented in Tanzania at a large scale, serves as a notable example, having contributed to the reduction of malaria transmission in this urban setting (24). Similarly, other countries have also prioritized larviciding; for instance, in Rwanda, drones are being used to implement larvicides in plantations (25), while in Malawi community-engagement plays a central role in larvicide application efforts (26). However, current evaluation methods for these interventions often rely on routine entomological monitoring and field observations, which can be resource-intensive, spatially limited, and slow to detect impacts.

Mathematical models are essential tools to guide stakeholders in making evidence-based decisions that are appropriate at a given scenario, the level of impact an intervention will bring, and predictions of future situations (27). However, many existing malaria models, including those of Kiware *et al*. which is Vector Control Optimization Model (VCOM), often rely on generalized parameters that may not capture local transmission dynamics, ecology, and seasonality. They use generalized scenarios simulating disease transmissions and impacts of interventions or predictions rather than local contexts that reflect the situationship and assist in making informed decisions. To address this gap, we aim to assess the impact of larviciding in combination with ITNs using a mathematical model based on district field-based data from Southeastern Tanzania, in malaria vector control, specifically *An. gambiae* s.l. in the Rufiji district as one of the dominant mosquito groups in the area. This will be achieved by using the field district-based data to inform the VCOM (28) model on the mortality rate of mosquitoes due to larviciding application. District-based modelling will ensure to capture variation in transmission ecology, seasonality impacts to the transmission and how different-intervention coverage levels impact malaria-vector control, providing evidence that is both rigorous and actionable for malaria control programs at that specific context.

## Methods

### Description of the study area

The dataset used in this study was generated during a pilot project implemented in the Rufiji district, found in the southeastern part of Tanzania at 7.8387° S, 38.3481° E. The Rufiji district covers 14, 500 km^2^ and the human activities in this area are mostly agriculture (rice) and fishing. The study took place in two wards that included 19 villages. The coverage of ITNs in this area is approximated to be 80% − 85% and the malaria prevalence of 26% [95% CI, 23.7-27.8] (29). There is no application of IRS in this area because in Tanzania IRS is applied in lake zone areas due to higher transmission in this zone. The Rufiji district can be seen from the Tanzania map in Fig 1

**Fig 1:**
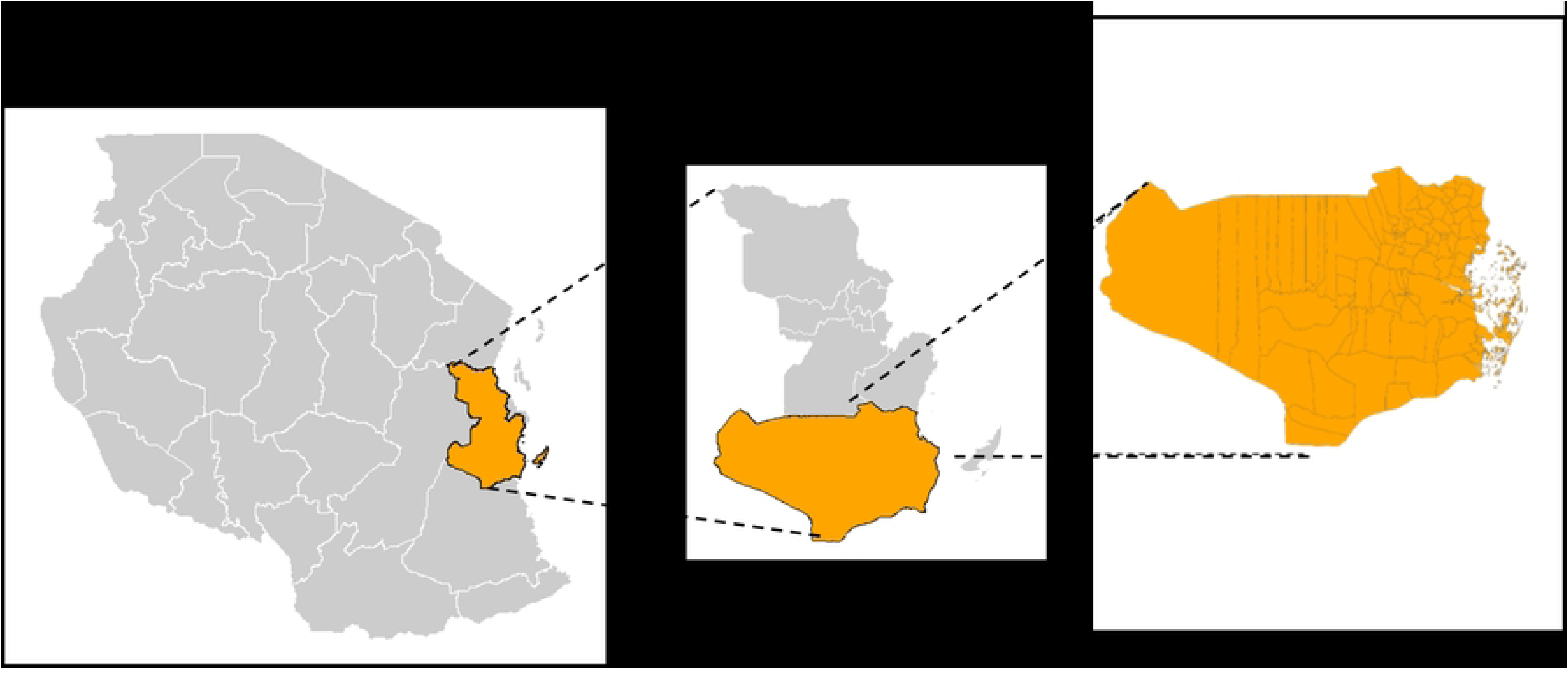
Project Implementation district

### Study design

Modelling components was not part of the original objective for this study done in Rufiji, the use of this data to answer the question of assessing the impact of larviciding makes it a retrospective study

### Data Collection

The project involved data collection during two phases, a baseline phase (2015–2016) where larviciding was not implemented in the Rufiji district, and an intervention phase (2020–2021) where larviciding was implemented. Both mature and immature mosquito population data were collected daily in the given areas of study. Other information collected with the number of mosquitoes includes the mosquito species, household identification number, location where the mosquito was collected if indoor or outdoor and the nature of the source in which the mosquito is collected if wet or dry. The malaria vectors were collected using three different methods: CDC-light traps, dipping, and clay pots. CDC-light traps were used to collect indoor matured mosquitoes while clay ports were used for both indoor and outdoor collection. Dipping was used for the collection of the immature mosquito population. Visualization of the monthly collected data of the female *An. gambiae* s.l. was done to show the visual difference in the monthly collections of the mosquitoes [fig4].

### Parameter Estimation

Both the first phase (control) and second phase (intervention) data were used to estimate the proportional increase in mortality of the mosquito population; the change in the mosquito density between the two phases stands as a proxy for the mortality estimated. Seasonality impact was accounted for when calibrating using a seasonal function (without meteorological effects) (30, 31) and the data fitted. The proportion effect of larviciding was obtained from a comparison between the two dataset parameters. Bayesian inference was performed in Stan (32) in Rstudio (33). For each parameterized model, we run four Markov chains with 2000 iterations, removing the first 500 for burn-in. The convergence of chains was checked using the diagnostics available within Stan, including the R-statistic and effective sample size. Hyperparameter posteriors were plotted as pairwise scatter plots to identify any potential parameter identifiability issues.

### Sensitivity analysis

Sensitivity analysis (34) is a method to determine how different dependent variables affect the output of the independent variable or model. For the parameters used in the VCOM model equations, sensitivity analysis was done to analyse how they drive malaria transmission in this scenario study. Knowing the sensitivity index of a parameter to the output will allow the control of the parameters that are responsible for increasing transmission (35). The values for the parameters and range variation that were used in the analysis can be seen in Table 1. The normalized forward sensitivity index method is used.

**Table 1:** Parameters used in the model and sensitivity analysis including their ranges.

| Parameter | Description | Value | Value Range | Source |
| --- | --- | --- | --- | --- |
| recRate | Human recovery rate | 0.15 | 0.05 - 0.2 | [21] |
| muVcom | Adult mosquito daily mortality in the presence of interventions | 0.315 | 0.1 - 0.5 | estimated |
| muPL | Pupal daily mortality | 0.25 | 0.2 - 0.3 | [21] |
| muLL | Late larval instar daily mortality | 0.035 | 0.03 - 0.05 | [21] |
| muEL | Early larval instar daily mortality | 0.034 | 0.02 - 0.05 | [21] |
| lambdaV | Force of infection in vectors at equilibrium | 0.01337 | 0.008 - 0.03 | [21] |
| durPL | Duration of pupal stage | 0.664 | 0.5 - 3 | [21] |
| durLL | Duration of late instar stage | 6.64 | 5 - 8 | [21] |
| durEL | Duration of early instar stage | 6.64 | 5 - 8 | [21] |
| durEV | Duration of latent period in mosquito (days) | 10 | 8 - 12 | [21] |
| Bh | transmission efficiency from an infectious mosquito to an uninfected, susceptible human | 0.5 | 0.4 - 0.6 | [21] |
| betaCom | Eggs laid per day by female mosquito in the presence of interventions | 21.19 | 15 - 25 | [21] |
| a_theta | human biting rate per mosquito in the presence of interventions | 0.3 | 0.05 - 0.5 | [21] |

### The Vector Control Optimization Model (VCOM)

The VCOM model (28) was adapted by applying the study area-specific parameters, which are mortality due to larviciding application and EIR for Rufiji. Other parameters from the literature and others originating from the VCOM model were used to study the impact of larviciding when added to ITNs [Table 1]. The model was adapted because it simulates mosquito life stages and their interactions with humans, in addition to incorporating various vector control interventions. This allows changes in the intervention coverage to demonstrate their impact on malaria vector control [fig 2]. The model was extended to capture the seasonality impact on malaria transmission (36). Several entomological outputs of the model can be considered as measures of the impact of intervention including, mosquito density, the reduction in human biting rate, and EIR (28).

**Fig 2:**
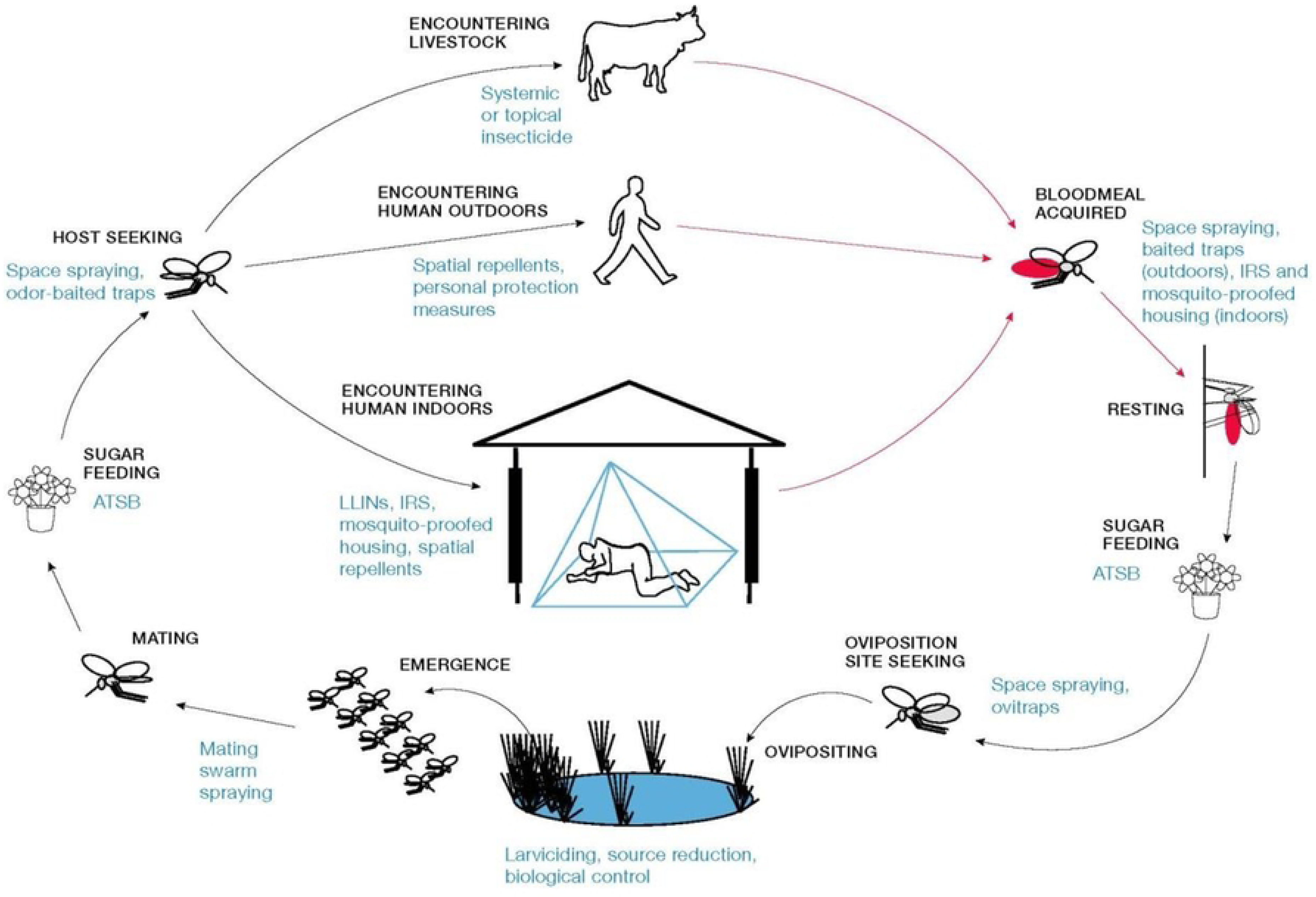
The schematic highlights opportunities for existing and novel vector control tools that can be used to target mosquitoes both indoors and outdoors and at all stages of the mosquito life and feeding cycles. Synergy and layering between interventions follow from this schematic diagram and how all the interventions are encoded from it (28).

A one-year simulation, modeled on a daily scale, was performed to examine the effects of different larviciding coverage levels in combination with varying ITNs coverage scenarios. Because not all interventions are implemented simultaneously, it was assumed that ITNs deployment would occur first, followed by larviciding. to reflect on real field situations. Each mosquito state was simulated to demonstrate the reductions achieved when these interventions are applied at different times or coverage levels. These simulations can be automatically viewed in the available software open access available in the <u>GUI</u> [http://skiware.github.io/VCOM/].

## Results

### Sensitivity analysis

Sensitivity analysis was conducted to evaluate the impact of various transmission parameters on the model’s output. Sensitivity indices were calculated for each parameter to quantify their influence on the model dynamics. The parameter definition and range are provided in Table 1. The results, illustrated in Fig 3, indicate that different parameters exhibit varying degrees of sensitivity. Specifically, the human recovery rate (recRate), duration of the latent period in mosquitos (durEV) and adult mosquito daily mortality in the presence of interventions (muVCom) have significant negative impacts, meaning that increases in these parameters lead to a reduction in transmission dynamics. Meanwhile, force of infection in vectors at equilibrium (lambdaV) and transmission efficiency from an infectious mosquito to an uninfected, susceptible human (bh) demonstrate a positive sensitivity index, suggesting that higher values of these parameters could potentially increase the transmission of malaria.

**Fig 3:**
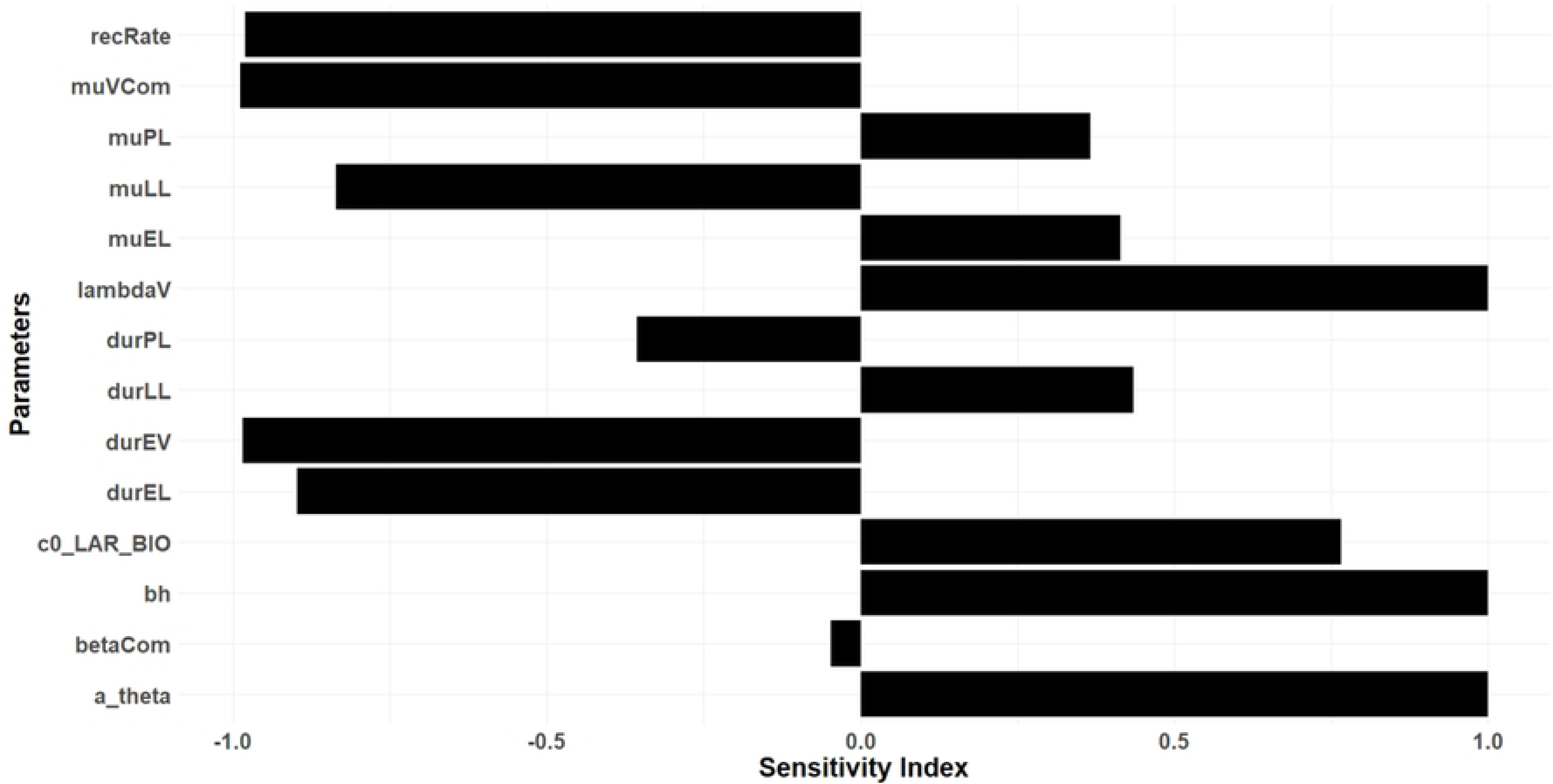
Sensitivity indexes indicating how parameters impact the transmission

### Effect of Larviciding on Mortality

Using the Bayesian inference approach, a proportion of 0.61 is obtained as an estimated impact to mortality. This value is directly used in the VCOM model as a proportional increase to the mortality value of the *An. gambiae* s.l species in the larval stage. The visualization of the two datasets, as shown in Fig 4, clearly illustrates this reduction.

**Fig 4:**
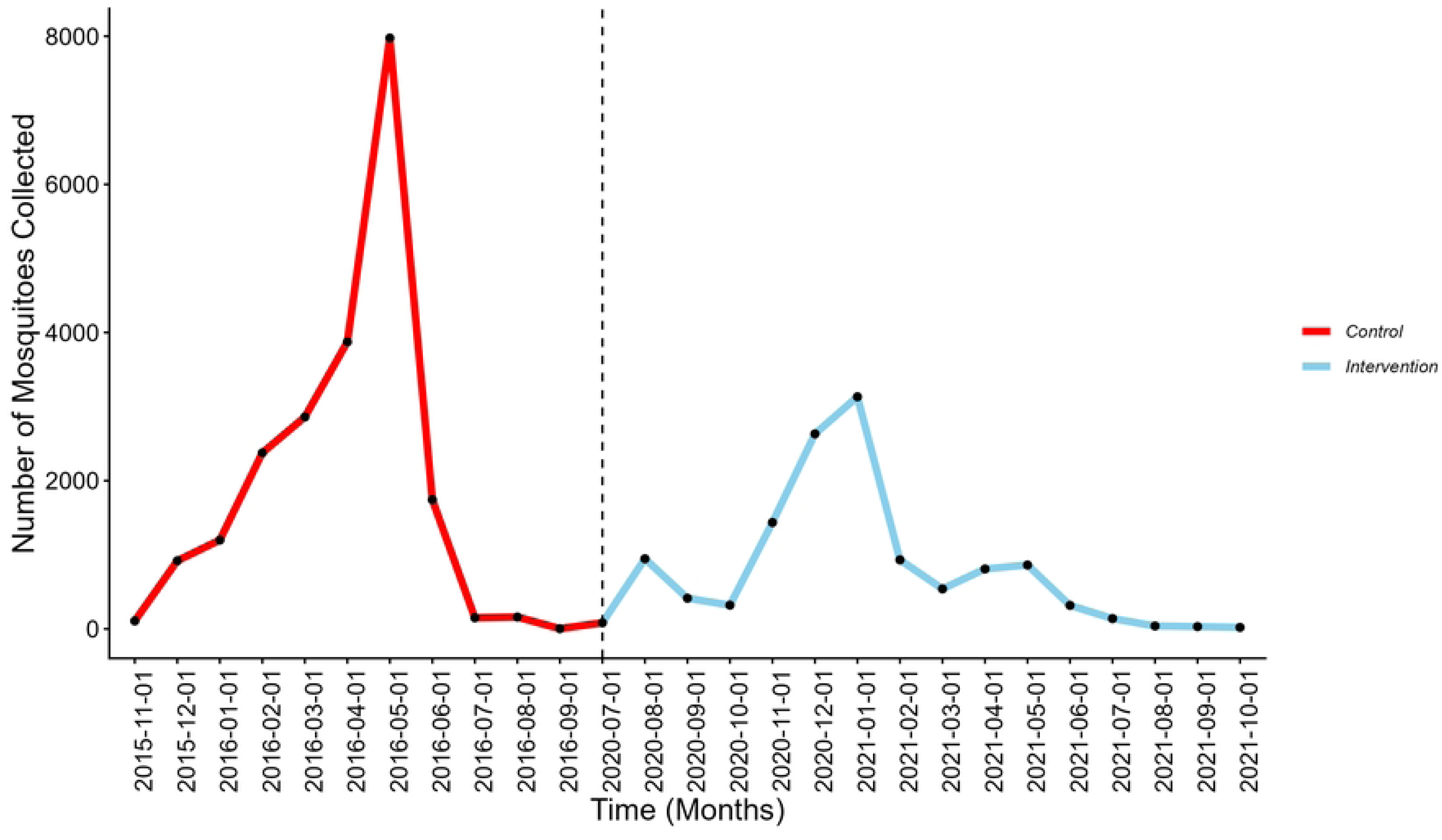
Monthly collection of Anopheles gambiae in the two phases

### Simulating the population

The results from the differential equations of the VCOM model were simulated according to mosquito populations and other entomological output measures. The simulation results for these two populations are in Fig 5. Because not all the interventions were implemented from the beginning, larviciding was introduced after ITNs. The simulation was then conducted, assuming the interventions were implemented at different times. Out of the 365 days in a year, the ITNs starts at the beginning, and larviciding on the 150th day.

**Fig 5.**
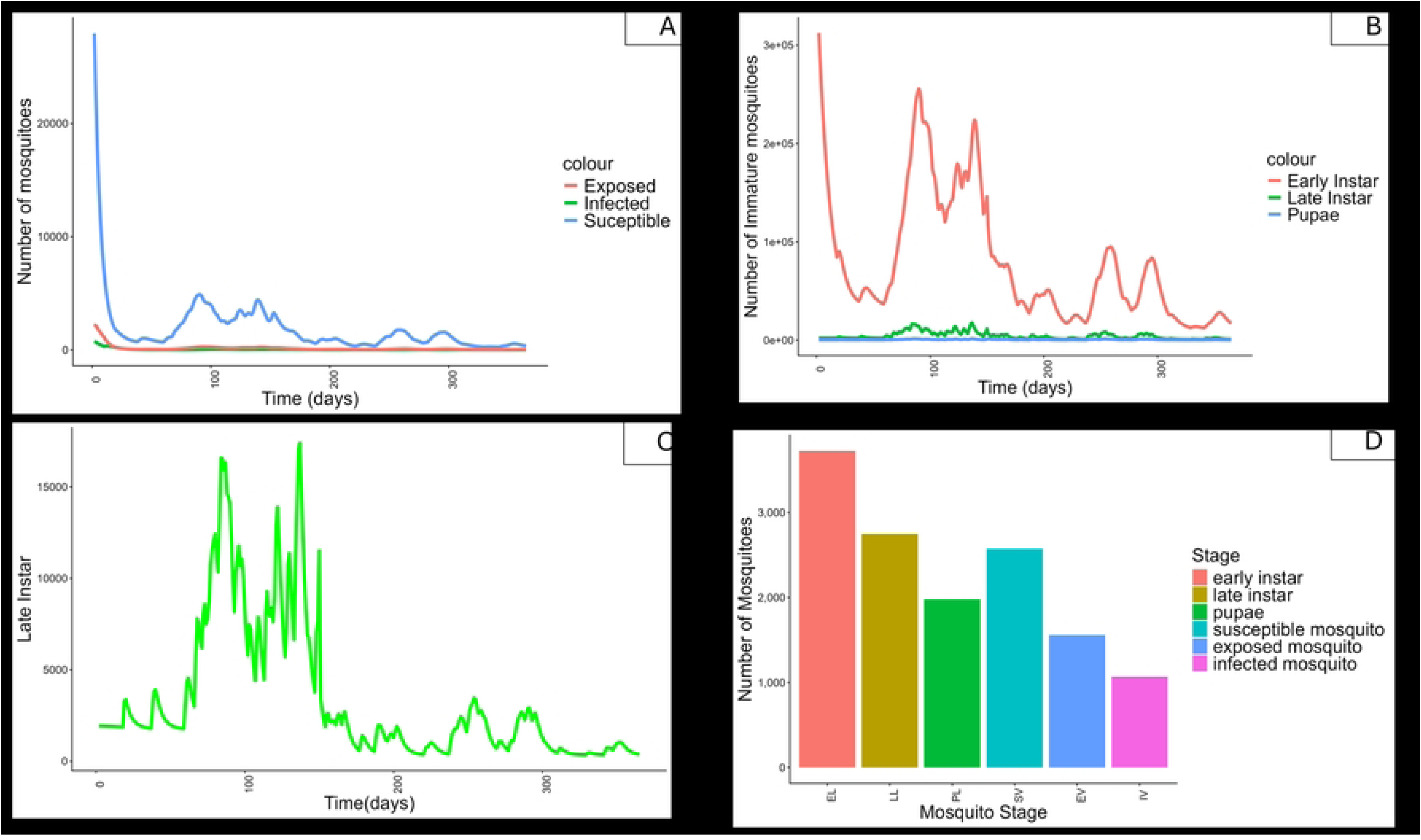
Simulation of the population involved in the model. (A) shows the matured mosquito population. (B) shows the immature population. (C) a zoomed version to display the late instar reduction in the application of these two interventions. (D) indicated the bar plot for total mosquito population.

All simulations were performed with the application of 80% ITNs and 60% larviciding to simulate these mosquito population dynamics. The mature mosquito population in Fig 5(a) indicates a vivid reduction in the susceptible population, while the infected and latent population also declines, though their numbers are small compared to the susceptible population. The small proportion of infected mosquitoes compared to the overall mosquito population is a result of the intervention’s impacts. For the immature mosquito population, the application of the intervention results in reducing the number of early larvae, along with other immature stages, as shown in Fig 5(b). Due to the impact of larviciding on the larval stage, a reduction in the number of late instar stages can be observed, as depicted in Fig 5(c), which is zoomed-in view from the previous Fig 5(b). A bar graph can also effectively visualize the total populations of the mosquito stages, revealing that the infected mosquito population remains very small for the given simulation period.

### Mosquito Density

Fig 6(b) shows that the mosquito-to-human ratio decreases more when two interventions are implemented compared to using a single intervention. A noticeable reduction in mosquito density is observed with the deployment of ITNs at 80% coverage (before larviciding) and even greater reduction when both ITNs at 80% coverage and larviciding at 60% coverage are implemented on the 150^th^ day of the year (after larviciding). The combined interventions result in a 43.9% decrease in mosquito density. This reduction is attributed to the effect of larvicides, which prevent immature mosquitoes from developing into adults, thereby directly reducing the overall mosquito population.

**Fig 6:**
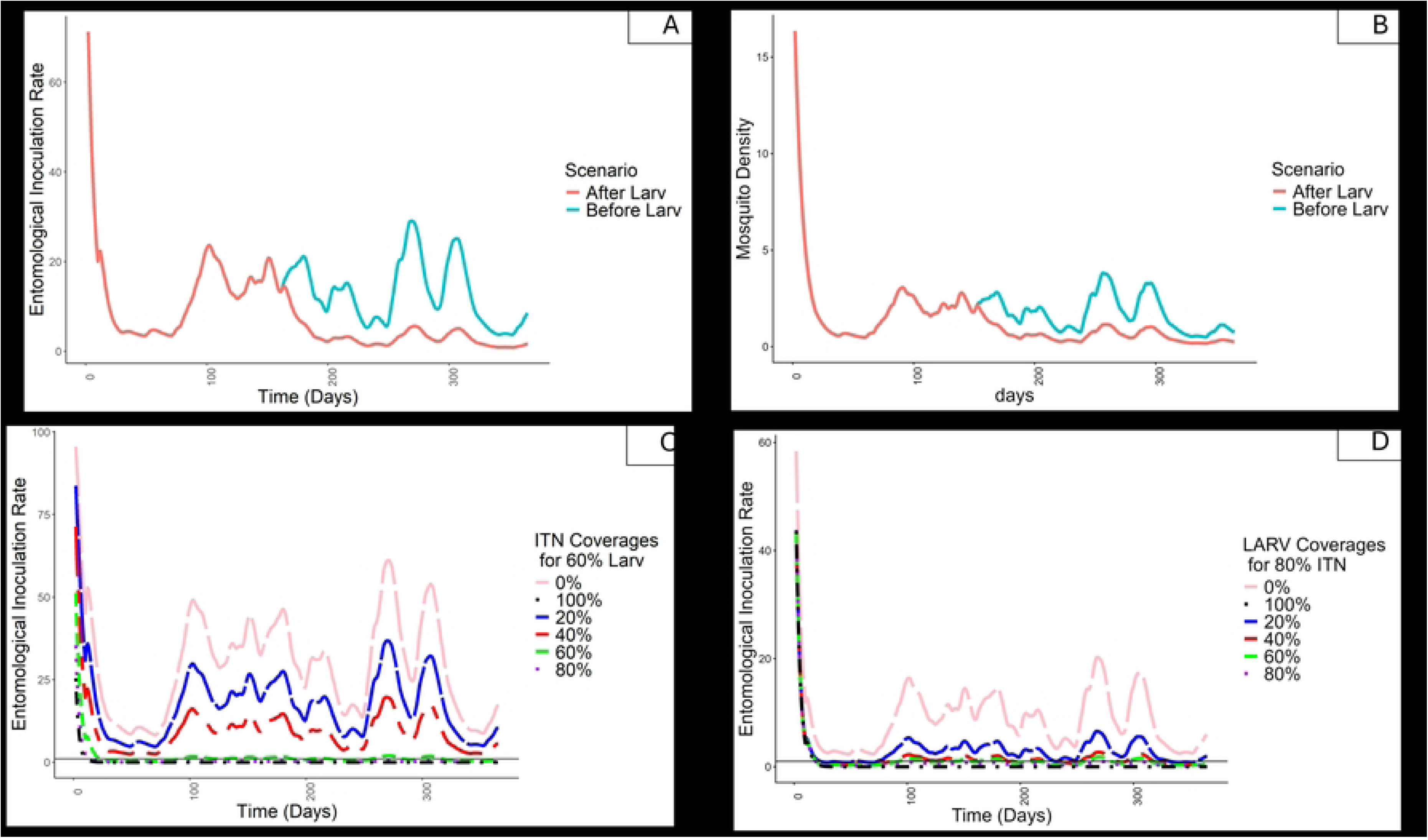
Simulated entomological outputs of the model. (A) Entomological inoculation rate (EIR) over time. (B) Adult mosquito density over time. (C) EIR under varying ITNs coverage with larviciding fixed at 60%. (D) EIR under varying larviciding coverage with ITNs fixed at 80%.

### Entomological Inoculation Rate

The value of EIR decreases over time following the implementation of interventions. EIR is measured in relation to the proportion of infectious mosquitoes within the population. As shown in Fig 6(a), a reduction in EIR is observed when ITNs are deployed at 80% coverage (before larviciding) and when both ITNs at 80% and larviciding at 60% coverage are implemented on the 150^th^ day of the year (after larviciding). The addition of larviciding results in a further 26.6% decrease in EIR. Increasing larviciding coverage in combination with ITNs leads to a reduction in EIR, allowing for analysis of different intervention scenarios’ output. The target is to achieve an EIR of less than one, which indicates interruption of malaria transmission. From Fig 6(d), this threshold is reached at larviciding coverages greater than 40%. Meanwhile, when only ITNs are used, the EIR does not fall below this threshold throughout the simulation period, as shown by the blue line in Fig 6(a). Although ITNs alone reduce EIR and lower transmission levels, residual transmission remains within the population. When varying ITNs coverage while maintaining larviciding at a constant 60%, Fig 6(c) shows that ITNs coverage above 60% can achieve an EIR below one.

## Discussion

This study evaluated the impact of incorporating larviciding alongside the existing ITNs intervention for malaria vector control in Rufiji District. The findings indicate that ITNs alone are insufficient to achieve optimal vector control. The entomological component of this work examined the effects of these interventions through a modeling framework, enabling a quantitative assessment of their combined impact on transmission dynamics. To the best of our knowledge, this represents the first study in Rufiji District to undertake a direct, model-based comparison assessing the additional benefit of larviciding on malaria vector populations. The study observed a reduction in the relative mosquito density of *An. gambiae s.l.,* following the application of larviciding in Rufiji, with an increase in mortality of 61%. From the simulation of the entomological outcomes, EIR was observed to drop by 22-fold in the use of both interventions and 9-fold in the use of only ITNs. A 7-fold drop in mosquito density was observed with the use of both interventions, while a 2-fold drop was observed with the use of only ITNs. This finding shows that additional complementary tools, such as larviciding, that ensure a reduction of mosquito density are essential.

There is considerable heterogeneity in the outcome measures used to assess the impact of larviciding intervention. Most studies have reported effect on the EIR, human biting exposure, and the larval or adult mosquito densities. In urban Dar es Salaam, a 96% reduction in *Anopheles* larvae was observed after one year of intervention, accompanied by a 21.3% reduction in EIR. Additionally, the intervention is highly recommended to be used as a supplementary intervention but not replacing the primary tools which are LLINs and IRS (37). The aspect of timing was discussed by Fillinger et al., who reported larviciding to reduced *Anopheles* larval density by 95% and human exposure to bites from adults by 92% in rural Kenya (9), and his other study in western Kenya that reported a >90% of larval density with 73% reduction in EIR (38). The intervention is said to be seasonal, with advice on the application of larviciding in the dry season and at the beginning of the rainy season. In Yaoundé, Cameroon it was observed with over a 68% reduction of *Anopheline* densities collected using CDC-LT and 79% reduction of EIR (39). In Rwanda reported Ano*pheles* reduction in the rice farms due to application of Larviciding in different rounds (40). In Burkina Faso using Bti based larviciding ensured the number of female *Anophelese* species to drop by 70% (19).

The application of larviciding has shown a positive impact on malaria vector control and is said to be a safe intervention with no adverse effects to other organisms (41). The IVM strategy, with involvement of larviciding as a supplementary tool will ensure reduction in malaria transmission by reducing the adult mosquito population. Moreover, the Bti used in this study to-date shows no resistance to its efficiency and is therefore highly advised (42). The limitation of this study includes, first, that attaining full larviciding coverage in a real-life scenario for all available habitats is difficult and requires extensive surveillance and community engagement. Nevertheless, the impact assessed by the model assumes proper implementation of larviciding to reach the required coverage, which is challenging in an operational field setting. This intervention is said to be applicable with an accurate measure of coverage in areas where mosquito habitats are few, fixed, and findable (16, 17). This argument is also discussed by Tusting et al., who suggest that larviciding can be applied successfully and at reasonable cost even in areas not traditionally recommended by WHO (43). To ensure maximization of coverage and efficiency, implementation should leverage innovative approaches such as the use of drones for precise mapping and targeted application of biological larvicides. Drones can enhance surveillance by detecting and mapping potential breeding sites that are difficult to identify through ground-based surveys, such as those formed in flooded fields, irrigation channels, or water-filled cow hoof prints common in livestock-keeping areas like Rufiji. By providing high-resolution imagery and geographic data, drone technology can help vector control teams target interventions more efficiently, reduce missed habitats, and improve resource allocation. Furthermore, the involvement of communities in larvicide application can complement these technologies, ensuring access to smaller or temporary habitats that may not be captured by aerial mapping, thereby expanding coverage and promoting sustainability (21). Secondly, the control data and intervention data were collected at two different time points; this may lead to differences in some external factors that cannot be captured in the model and so inconsistencies. Thirdly, the issue of community involvement is essential in LSM and larviciding itself, this factor was not mostly considered in this modeling study.

However, this study has demonstrated the additional advantage to the fight against malaria vector control that can be attained when larviciding is used in combination with ITNs. The effectiveness observed when larviciding is deployed complementing ITNs use ensures more interruption in malaria transmission in the area. Future study should consider impact assessment for the larval source management intervention including the quantification of source management component, this will require study to collect the data on how the SM was performed and its contribution to vector population, this will inform the model. However, the impact of this intervention beyond malaria vector species can be assessed in the future.

## Conclusion

This study provides valuable insights into the potential of larviciding as an effective and key strategy for malaria control in the local context. While promising results were observed, the study also highlights the operational challenges of achieving optimal larviciding coverage as suggested by the model outcomes. Innovative solutions such as the use of drones could enhance the identification and mapping of mosquito breeding habitats, particularly in hard-to-reach or inaccessible areas, including those associated with livestock activity such as puddles formed by cow hooves in Rufiji. Strengthening community engagement will further support sustainability and ensure wider and more effective larviciding coverage. Overall, these findings contribute to the broader understanding of integrated vector control strategies and provide a foundation for future studies and policy development.

## Data Availability

The datasets used and/or analyzed during the current study are available in an open-source database (mosquitodb.io), the corresponding author will give access to the specific data upon reasonable request.

## Acknowledgement

We would like to express our sincere gratitude to the technicians, data collectors, volunteers, villagers where the study was conducted and organizations that generously provided access to their resources, which greatly contributed to the development of this manuscript. Their insights have significantly enhanced the quality of our work.

